# Rare coding variation in OCD implicates shared genes with other psychiatric disorders

**DOI:** 10.64898/2025.12.22.25342827

**Authors:** Seulgi Jung, Matthew W. Halvorson, Nancy Pedersen, Marina Natividad Avila, Maria Niarchou, EGOS, NORDiC, Bernie Devlin, Kathryn Roeder, James J. Crowley, Joseph D. Buxbaum, Dorothy E. Grice

## Abstract

**Importance:** Obsessive-compulsive disorder (OCD) affects 2-3% of the population with often disabling obsessions and compulsions. Despite its high heritability, genetic studies of OCD have lagged other psychiatric disorders, particularly in understanding the role of rare genetic variants.

**Objective:** To identify rare coding genetic variants contributing to OCD risk and examine genetic overlap with chronic tic disorders (CTD) and other psychiatric conditions.

**Design:** Family-based and case-control whole-exome sequencing (WES) study.

**Settings:** WES data were aggregated from 11 independent cohorts across Sweden, the United States, and the United Kingdom.

**Participants:** A total of 47,194 individuals were available, and 44,089 passed quality control for analysis. The final sample included 6,071 individuals with OCD, comprising 1,202 probands from family-based trios and 4,869 cases, and 38,018 controls.

**Exposures:** Rare damaging coding variants identified by WES.

**Main Outcomes and Measures:** Identification of OCD risk genes through rare variant analyses, meta-analysis with CTD data, gene-set enrichment analyses, and evaluation of cross-disorder genetic overlap using curated gene sets.

**Results:** The analysis provided an estimate of approximately 470 autosomal genes contributing to OCD risk through rare genetic variation. *CHD8* reached genome-wide significance (q < 0.05). Meta-analysis with CTD data revealed additional risk genes, including *CELSR3* (q < 0.05), *QRICH1*, and *WWC1* (q < 0.1). We observed significant genetic overlap between OCD, autism spectrum disorder (ASD), and developmental delay: 33% of ASD genes with FDR < 0.1 showed association with OCD (p < 0.001), and 36% showed possible associations in the shared OCD-CTD genetic architecture (p < 0.001), but minimal rare-variant overlap with bipolar disorder and schizophrenia risk genes. We also found that *CHD8-*regulated genes were enriched for both rare and common variant associations with OCD.

**Conclusions and Relevance:** In this largest study to date of rare coding variation in OCD, we confirm *CHD8* as the first genome-wide significant rare-variant risk gene, show that genes that are targets of *CHD8* can carry rare and common variant risk for OCD, and identify multiple additional genes and pathways contributing to risk. Taken together, the findings show that OCD shares substantially greater genetic overlap with neurodevelopmental conditions than with adult-onset psychiatric disorders, refining the developmental framework of OCD and informing future mechanistic and clinical research.

**Key Points:** *Question:* How do rare genetic variants contribute to OCD, and how do they overlap with variants linked to chronic tic disorders (CTD) and other neurodevelopmental conditions?

*Findings:* Exome sequencing of 6,071 OCD cases demonstrated significant enrichment of rare damaging variants in evolutionarily conserved genes, and *CHD8* emerged as the first genome-wide significant OCD risk gene. Rare variant patterns in OCD and CTD aligned with those seen in neurodevelopmental, but not adult-onset psychiatric disorders, indicating shared neurodevelopmental pathways.

*Meaning:* These findings clarify OCD’s neurodevelopmental genetic architecture, identify *CHD8* as a key risk pathway, and reveal overlap with CTD and other conditions.

## Introduction

Obsessive-compulsive disorder (OCD) has a lifetime prevalence of 2-3% and is characterized by intrusive and unwanted thoughts, images, or urges (obsessions), and repetitive behaviors or mental rituals (compulsions) that typically function to reduce the distress associated with obsessions. OCD has a substantial genetic component, with heritability estimates ranging from 35 to 50%.^1–11^ While much of this heritability is attributed to common genetic variation,^7,12–14^ rare genetic variants, including copy number variants (CNVs), single nucleotide variants (SNVs) and small insertions and deletions (indels), also confer risk for OCD.^15–18^ However, prior OCD sequencing studies have been underpowered, yielding suggestive but not genome-wide significant risk genes. Despite their potential to shed light on OCD pathobiology and identify therapeutic targets, rare variants remain poorly studied in OCD compared to other psychiatric disorders, where large-scale sequencing studies have revealed high-confidence risk genes.^16,17,19–23^

OCD frequently co-occurs with chronic tic disorders (CTD), including Tourette syndrome and persistent motor or vocal tic disorders.^24^ Individuals with OCD are at elevated risk for CTD, and individuals with CTD are at an elevated risk for OCD, with shared genetic risk between the two conditions well established.^25–31^ Rare variants have also been implicated in CTD,^15^ though, as with OCD, they remain poorly studied due to the limited availability of sufficiently powered sequencing datasets. Meta-analysis of rare variants in OCD and CTD provides a unique opportunity to identify shared genetic pathways, enabling the discovery of biological pathways underlying both disorders and revealing shared mechanisms that can inform cross-disorder treatment.

To address these gaps, we generated novel exome sequencing data from 1,665 OCD cases and 450 controls from the Epidemiology and Genetics of Obsessive-Compulsive Disorder in Sweden (EGOS) cohort.^11,32^ We integrated these data with exome sequences from 105 OCD cases and 521 matched controls in the Mount Sinai BioMe biobank,^33,34^ and additional previously sequenced OCD cohorts. The final sample included 6,071 individuals with OCD, comprising 1,202 probands from family-based trios and an additional 4,869 cases and 38,018 controls, representing a dataset approximately six times larger than any previous rare-variant study of OCD. Leveraging this unprecedented sample, we analyzed rare coding variants associated with OCD and performed a meta-analysis incorporating OCD and CTD sequencing data to identify genes influencing both disorders.

## Methods

### Data Source and Study Population

A total of 47,194 samples were aggregated from 11 independent sources: (1) The EGOS collection from Sweden (n= 2,115: 1,665 OCD cases and 450 controls);^32^ (2) LifeGene Swedish controls were sourced from the LifeGene Biorepository at the Karolinska Institute (n= 4,765);^35^ (3) The Mount Sinai BioMe Biobank, founded in 2007, is an ongoing electronic health record-linked biorepository of the Mount Sinai Health System, comprising approximately 60,000 participants.^33,34^ We used gVCF files of 105 OCD cases and 525 controls without mental disorders (n= 630); (4) Yale (n= 666: 222 trios);^17^ (5) Columbia/Johns Hopkins (n= 4,245: 663 trios, 495 cases, and 1,761 controls);^16^ (6) All of Us database (https://allofus.nih.gov/) (n= 9,654: 1,609 cases and 8,045 controls without mental disorders);^36^ (7) UK Biobank (https://www.ukbiobank.ac.uk/) (n= 7,518: of 1,253 cases and 6,265 controls without mental disorders);^37^ (8) Simons Foundation Powering Autism Research for Knowledge (SPARK; n = 156: 52 trios; the probands in these trios were not diagnosed with autism);^38^ (9) Lin et al., 2022 (n = 159: 53 trios);^39^ (10) Wang et al., 2018 (n= 918: 306 trios);^26^ (11) Fu et al., 2022 (n=16,368:5,456 unaffected sibling trios).^40^ The distribution of these cohorts is provided in eTable 1 in Supplement 2.

### Genetic Data Analysis

The data included both family-based and case-control designs, with samples processed through whole-exome sequencing (WES). The sequencing data were aligned to the human reference genome (GRCh38) using bwa v.0.7.15^41^ and processed with the Genome Analysis Toolkit (GATK)^42^ to generate gVCFs, which were subsequently combined for joint genotyping. Extensive quality control (QC) measures were implemented, including trimming and alignment of sequencing reads, duplicate marking, recalibration of base quality scores, and variant filtering. These steps ensured high data integrity across cohorts, with samples subjected to contamination and relatedness checks using Hail 0.2 (https://hail.is) and PRIMUS.^43^ Variants were annotated with the Ensembl Variant Effect Predictor (VEP) to assess their functional impacts.^44^ Details of joint genotyping and QC are provided in the eMethods in Supplement 1.

De novo variants were identified using Hail’s de_novo() function, with strict thresholds for allele balance, read depth, and prior population allele frequencies from gnomAD (version 3.1.2).^45^ We kept de novo variants with an allele frequency ≤ 0.1% in our dataset, and in the non-psychiatric subset of the gnomAD database (version 3.1.2).^45^ Rare inherited variants were identified using the transmission disequilibrium test (TDT) in Hail. For case-control analyses, principal components analysis (PCA) was used to verify ancestry matching, and cohorts were matched on sex and ancestry using the “optmatch” R package. The TDT and case-control variants were required to have an allele count of ≤5 in our dataset, and in the non-psychiatric subset of the gnomAD database (version 3.1.2).^45^

Copy number variants (CNVs) were detected using GATK-gCNV,^46^ employing read depth normalization and principal component analysis to account for batch effects. High-confidence CNVs were annotated against curated genomic disorder loci and classified based on their overlap with gene exons. De novo CNVs were identified by comparing offspring CNVs with parental genomes, ensuring high-quality thresholds for detection (see eMethods in Supplement 1).

### Statistical Analysis

The study utilized the Transmission and De Novo Association (TADA) model to integrate de novo, inherited, and case-control variant data for gene-level analyses.^47^ TADA employs a Bayesian framework to calculate Bayes factors (BFs) and false discovery rates (FDRs), enabling the identification of candidate risk genes. We calculated the q value for FDR. Genes with an q value less than 0.1 were classified as “high-confidence” risk genes, whereas those with an q value below 0.3 were designated as “potential” risk genes, thresholds applied in rare variant studies.^16,17,48,49^ Variants were grouped into protein-truncating variants (PTVs), missense variants classified by the ‘missense badness, PolyPhen-2, and constraint’ (MPC) scores,^50^ and synonymous variants. For CNVs, only those affecting constrained genes with low loss-of-function tolerance were included (see eMethods in Supplement 1).

To identify shared risk factors between OCD and CTD, we conducted a meta-analysis of WES data. Using TADA, gene-level BFs were calculated separately for OCD, based on our merged WES data, and for CTD, using data from existing studies, for each variant class. To identify genes contributing to both OCD and CTD, we multiplied the Bayes factors from the two datasets. This approach combined independent evidence of genetic association from both disorders, ensuring that genes with consistent signals in OCD and CTD were prioritized as shared risk factors. Finally, q values were calculated to account for multiple testing. Enrichment analyses assessed the overlap of OCD risk genes with curated gene sets, including pathway genes of *CHD8* and *ZMYM2*, as well as genes implicated in ASD, developmental delay, bipolar disorder, and schizophrenia. Permutation tests were used to evaluate statistical significance, while MAGMA (v1.10)^51^ was employed for gene-set analyses using genome-wide association study (GWAS) data (see eMethods in Supplement 1).

## Results

### Patterns of rare coding variants in OCD

To identify OCD risk genes impacted by rare genetic variation, we analyzed sequence data from 11 distinct cohorts (eTable 1 in Supplement 2). After quality control, these cohorts collectively included a total of 44,089 individuals, of whom 6,071 were diagnosed with OCD and others were parental or ancestry-matched controls.

The average number of rare de novo coding variants observed in OCD probands was notably higher compared to that in unaffected individuals (eTable 2 in Supplement 2). Specifically, OCD probands exhibited an average of 1.88 rare de novo coding variants per sample (2,255 variants in 1,202 probands) compared to 1.64 per sample in unaffected individuals (8,982 variants in 5,456 individuals). This difference was statistically significant (*P* = 4.47E-08, binomial test).

To delve deeper into the sources of these elevated variant numbers, we categorized the variants into PTVs and missense variants (Figure 1). Counts of de novo PTVs in OCD probands were significantly higher than those in unaffected individuals (0.11 in probands, compared to 0.09 in unaffected siblings; *P* = 0.03, binomial test). We further analyzed the genes by dividing them into deciles based on the ‘loss-of-function observed/expected upper bound fraction’ (LOEUF) score,^52^ an empirical measure of gene-level evolutionary constraint. Our analyses indicated that the signal for de novo PTV was concentrated in the most conserved genes and this pattern was similarly observed for PTVs in cases versus controls (Figure 1).

**Figure 1.**
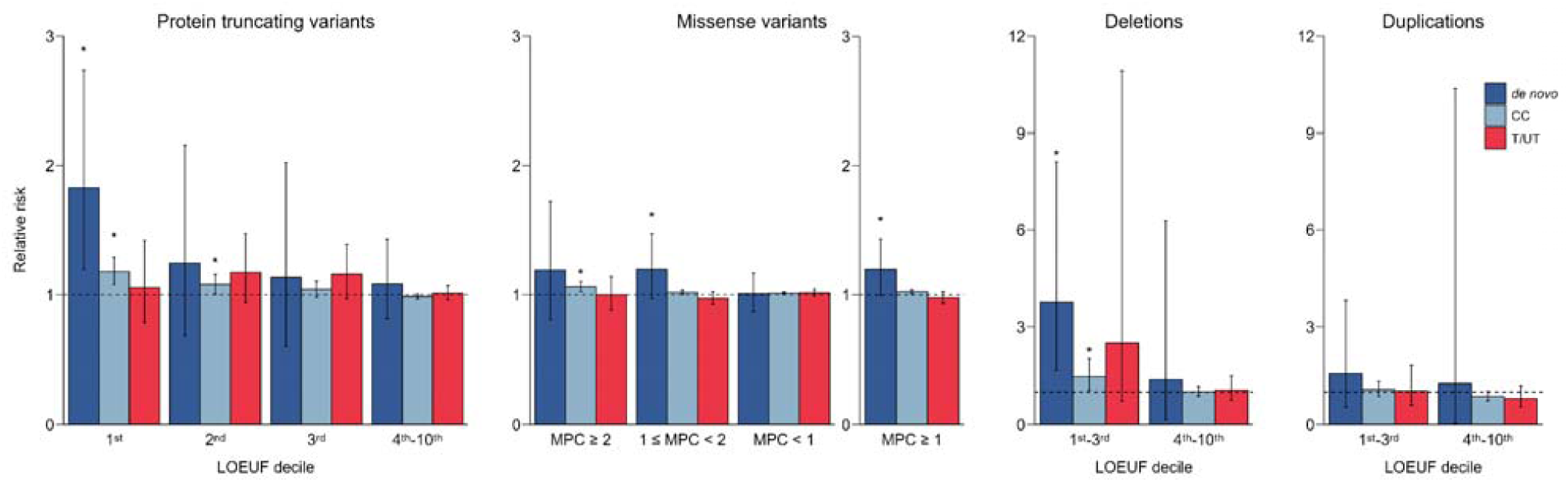
Rates of single nucleotide and copy number variants. Relative rates of protein truncating variants (PTVs), missense variants in constrained genes, and deletions and duplications of genic copy variants (gCNVs) are shown. For each, relative rates for de novo variation, case control variation, and inherited variation (as transmitted/non-transmitted variation) are shown. PTV and gCNV are subsetted by deciles of the LOEUF (Loss-of-function observed/expected upper bound fraction) score. For missense variants genes in the first-third deciles of LOEUF were subsetted using MPC (Missense badness, PolyPhen-2, and Constraint) scores as shown. \**P*<0.05, in binomial tests.

When examining missense variants, we categorized them into three sub-groups based on the estimated impact of the mutation using the MPC score.^50^ These categories were MisB (MPC ≥ 2), MisA (2 > MPC ≥ 1), and all other missense variants. We observed a significant signal for de novo MisA and MisB variants in OCD risk, particularly in genes with a LOEUF score within the first three deciles (Figure 1).

We next examined genic copy number variants (gCNVs) by applying the GATK-gCNV tool^46^ to the sequencing data, including external data where possible (eTable 3 in Supplement 2). We focused on small gCNVs harboring constrained genes, limiting our analysis to gCNVs that included eight or fewer genes and assigning the minimum LOEUF score across the genes to each gCNV. Our analysis revealed significant enrichments of both de novo (relative risk = 3.77, *P*=9.26E-04, binomial test) and case-control (relative risk = 1.46, *P*=1.85E-02, binomial test) deletions that included constrained genes, particularly those within the first three deciles of LOEUF (Figure 1; eTable 4 in Supplement 2).

### Gene discovery in OCD and CTD

To estimate the number of autosomal genes that can contribute to OCD risk when mutated, we employed a method developed by He and colleagues.^40,47^ Our analysis estimated that approximately 470 autosomal coding genes can contribute to OCD risk through rare deleterious variation, which corresponds to 2.60% of the genes analyzed (95% CI: 2.56-2.63%). These results suggest that OCD risk from rare deleterious variants is distributed across a substantial number of genes.

For gene discovery, we utilized an improved Transmitted and De Novo Association (TADA) method.^40,47^ TADA requires an estimate of the fraction of autosomal coding genes that can contribute to OCD risk through rare deleterious variation (approximately 2.60% from the above analysis) and an estimate of the average relative risk of variants as a function of gene constraint, BF and the q value (FDR) per gene (eTable 5 in Supplement 2).

Our TADA analysis identified *CHD8* as a genome-wide significant OCD risk gene (q value = 0.038) (Table 1 and Figure 2). Additionally, three other likely OCD risk genes were identified with q values less than 0.3, including *HECTD4*, *EML5*, and *SCUBE1*. Given the well-established evidence for clinical and genetic overlap between OCD and CTD^17,24,26,29,30,53,54^ we then performed a joint analysis of our OCD data with published CTD data (n=523 complete trios), specifically rare de novo coding variant counts from three CTD studies (eTable 1 in Supplement 2). From the combined meta-analysis of OCD and CTD data (eTable 6 in Supplement 2), two genes (*CELSR3* and *CHD8*) showed genome-wide significance. Furthermore, two additional genes (*WWC1* and *QRICH1*) had q values less than 0.1, and a total of 17 genes had q values less than 0.3 (Table 1 and Figure 2).

**Figure 2.**
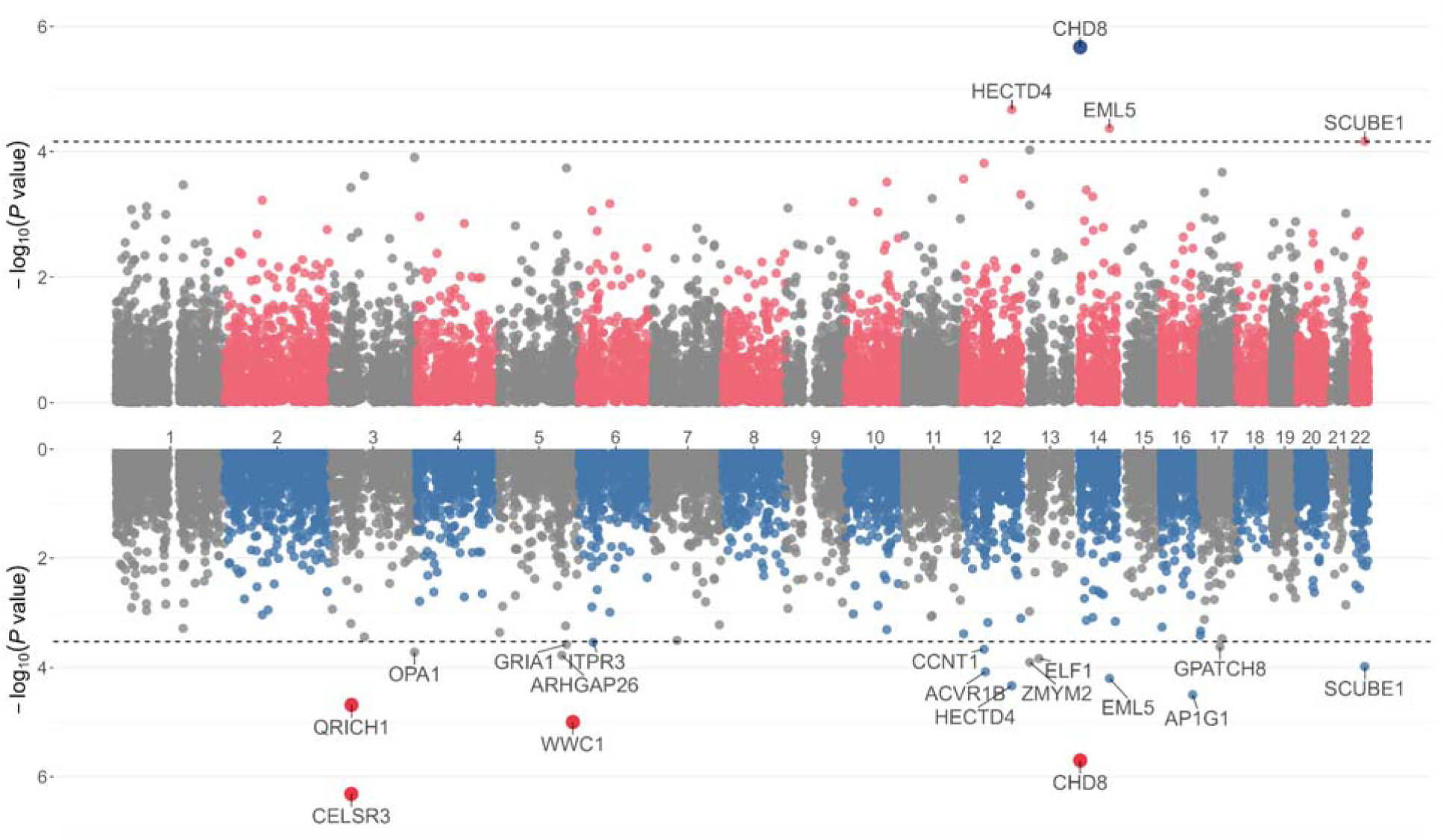
Identification of risk genes in OCD and CTD. *Upper,* Manhattan plot of TADA analysis for OCD. *Lower,* Manhattan plot of meta-analysis of the meta-analysis of OCD and CTD. Each dot symbolizes an individual gene. Genes that exhibit a q-value less than 0.1 are highlighted in blue (upper) or red (lower). Genes with q value <0.3 (dotted lines) are named.

**Table 1.**
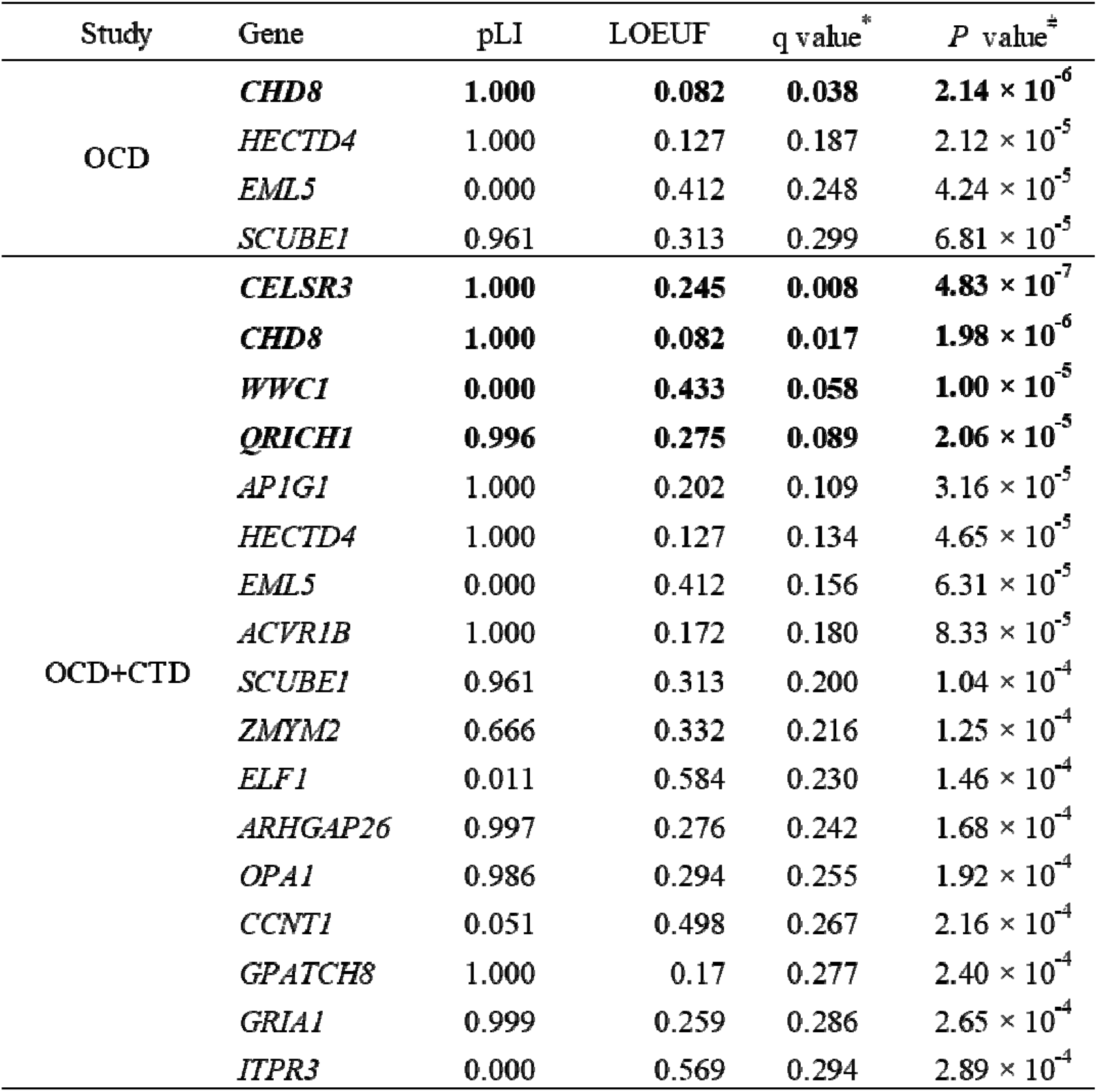
Top gene findings (q<0.3) in OCD and in the meta-analysis of OCD and CTD. Results for genes with q < 0.3 are shown, ranked by q value, with q value < 0.1 highlighted in bold text. For each gene, the q_value (false discovery rate) and *P* value (both calculated from the transmission and de novo association test) are shown, as are two metrics of gene-level constraint, pLI and LOEUF. CTD, chronic tic disorder; LOEUF, loss-of-function observed/expected upper bound fraction; OCD, obsessive-compulsive disorder; pLI, probability of loss-of-function intolerant.

| Study | Gene | pLI | LOEUF | q value* | P value<sup>‡</sup> |
| --- | --- | --- | --- | --- | --- |
| OCD | **CHD8** | **1.000** | **0.082** | **0.038** | **2.14 × 10<sup>-6</sup>** |
|  | *HECTD4* | 1.000 | 0.127 | 0.187 | 2.12 × 10<sup>-5</sup> |
|  | *EML5* | 0.000 | 0.412 | 0.248 | 4.24 × 10<sup>-5</sup> |
|  | *SCUBE1* | 0.961 | 0.313 | 0.299 | 6.81 × 10<sup>-5</sup> |
| OCD+CTD | **CELSR3** | **1.000** | **0.245** | **0.008** | **4.83 × 10<sup>-7</sup>** |
|  | **CHD8** | **1.000** | **0.082** | **0.017** | **1.98 × 10<sup>-6</sup>** |
|  | *WWC1* | **0.000** | **0.433** | **0.058** | **1.00 × 10<sup>-5</sup>** |
|  | **QRICH1** | **0.996** | **0.275** | **0.089** | **2.06 × 10<sup>-5</sup>** |
|  | *AP1G1* | 1.000 | 0.202 | 0.109 | 3.16 × 10<sup>-5</sup> |
|  | *HECTD4* | 1.000 | 0.127 | 0.134 | 4.65 × 10<sup>-5</sup> |
|  | *EML5* | 0.000 | 0.412 | 0.156 | 6.31 × 10<sup>-5</sup> |
|  | *ACVR1B* | 1.000 | 0.172 | 0.180 | 8.33 × 10<sup>-5</sup> |
|  | *SCUBE1* | 0.961 | 0.313 | 0.200 | 1.04 × 10<sup>-4</sup> |
|  | *ZMYM2* | 0.666 | 0.332 | 0.216 | 1.25 × 10<sup>-4</sup> |
|  | *ELF1* | 0.011 | 0.584 | 0.230 | 1.46 × 10<sup>-4</sup> |
|  | *ARHGAP26* | 0.997 | 0.276 | 0.242 | 1.68 × 10<sup>-4</sup> |
|  | *OPA1* | 0.986 | 0.294 | 0.255 | 1.92 × 10<sup>-4</sup> |
|  | *CCNT1* | 0.051 | 0.498 | 0.267 | 2.16 × 10<sup>-4</sup> |
|  | *GPATCH8* | 1.000 | 0.17 | 0.277 | 2.40 × 10<sup>-4</sup> |
|  | *GRIA1* | 0.999 | 0.259 | 0.286 | 2.65 × 10<sup>-4</sup> |
|  | *ITPR3* | 0.000 | 0.569 | 0.294 | 2.89 × 10<sup>-4</sup> |

### Cross disorder risk for OCD and CTD genes with other psychiatric disorders

To explore the overlap of genetic findings in OCD and CTD with other psychiatric and developmental disorders, we compared our results with published findings for ASD and developmental delay (DD) (eTable 7 in Supplement 2).^40^ Among the top four genes (q < 0.3) from the OCD analyses, mutations in two genes (*CHD8* and *HECTD4*) showed significant association with ASD and DD (eFigure 1 in Supplement 1). Additionally, mutations in four of the top (q < 0.3) 17 genes (*CHD8*, *HECTD4*, *ZMYM2*, and *ITPR3*) from the meta-analysis of OCD and CTD showed significant association with ASD (eFigure 1 in Supplement 1). *QRICH1* showed a q value of 0.063 for ASD and genome-wide significance for DD (eFigure 1 in Supplement 1). *AP1G1* was also a significant finding in DD.

Given this overlap, we examined the proportion of ASD genes from Fu et al.^40^ that showed evidence of association in our dataset.^55,56^ We observed that 33% of the 255 ASD genes with FDR < 0.1 from Fu et al. were associated with OCD in our analyses (P < 0.001), while 36% of the ASD genes showed possible associations in the joint analysis of OCD and CTD (P < 0.001) (Table 2). This significant enrichment indicates that the rare-variant architecture of OCD and CTD overlaps substantially with that of ASD, reinforcing a shared neurodevelopmental genetic basis across these conditions.

**Table 2.**
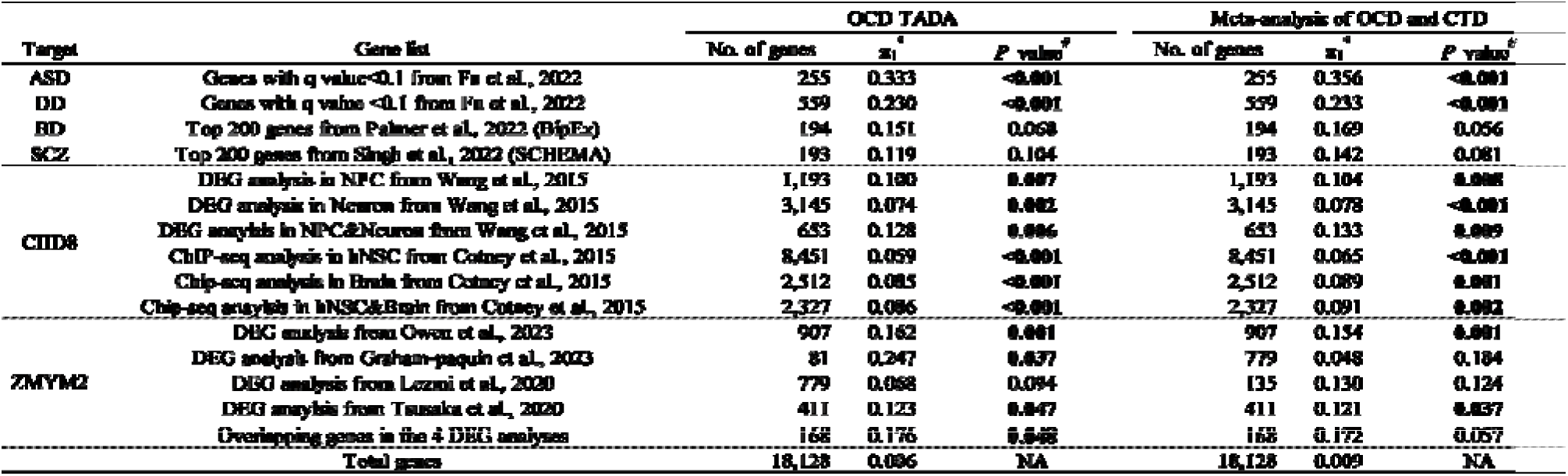
OCD and the meta-analysis of OCD and CTD genes overlap with other psychiatri disorders and include genes regulated by select transcription factors. . Gene lists from the indicated studies were assessed for evidence of association with OCD and the meta-analysis of OCD and CTD genes. For *CHD8* we also created lists that represented the intersection of other lists from the same study, while for *ZMYM2*, we created a list that included all genes found in 2 or more studies (n=168). For each list, the number of genes analyzed is shown as is π1 (proportion of genes showing deviation from the null) and the associated permutation-based *P* values. Gene lists with *P* value < 0.05 are bolded. ASD, autism spectrum disorder; BD, bipolar disorder; BipEx, the bipolar exome sequencing project (Palmer et al., 2022); *CHD8*, Chromodomain-helicase-DNA-binding protein 8; Cotney_brain, *CHD8* pathway gene list identified in human midfetal brain cells (Cotney et al., 2015); Cotney_hNSC, *CHD8* pathway gene list identified in human neural stem cells (Cotney et al., 2015); CTD, chronic tic disorder; DEG, differentially expressed genes; NA, not applicable; No. of genes, number of genes overlapped with gene-list of TADA result; OCD, obsessive-compulsive disorder; SCHEMA, the schizophrenia exome sequencing meta-analysis consortium (Singh et al., 2022); SCZ, schizophrenia; Wang_Neuron, *CHD8* pathway gene list identified in human early differentiating neurons (Wang et al., 2015); Wang_NPC, *CHD8* pathway gene list identified in human neural progenitor cells (Wang et al., 2015).

To verify that these estimates were not biased upward due to violations of model assumptions, we also estimated the proportion of genes showing evidence across the entire set of genes. Among all 18,128 genes, we estimated that 0.56% showed evidence of association with OCD alone, and 0.92% with the meta-analysis of OCD and CTD.

Recent large-scale analyses of rare variants have been reported for both bipolar disorder and schizophrenia.^45,57^ We examined the top 200 genes identified in these studies (Table 2). However, we did not observe substantial overlap with bipolar or schizophrenia risk genes. Interestingly, four genes (*CHD8, QRICH1, HECTD4,* and *ZMYM2)* with comparatively lower *P* values in the meta-analysis of OCD and CTD were implicated in three or more disorders in the above analyses (eFigure 3 and eFigure 1 in Supplement 1).^40,45,57–61^ The minimal overlap with BD and SCZ suggests that rare coding variation in OCD and CTD aligns more closely with neurodevelopmental disorders than with adult-onset psychiatric conditions. However, these results should also be interpreted with caution, given the limited power of these studies.

### Role of *CHD8* and *ZMYM2* pathway genes in OCD and CTD risk

*CHD8* is a chromatin remodeling gene previously implicated in the risk of ASD and DD. ASD and OCD frequently co-occur, and genes regulated by *CHD8* have also been associated with ASD risk.^15,62–65^ Furthermore, *CHD8* pathway genes have been implicated in bipolar disorder and schizophrenia,^45,65^ both of which are associated with an increased risk of OCD.^15,66–68^ Based on this evidence, we investigated whether *CHD8* pathway genes,^64,65^ identified through binding site proximity and differential gene expression analyses, were enriched for rare variants in our datasets. We found that up to 13% of *CHD8* pathway genes showed evidence of association with OCD or with the shared genetic basis of OCD and CTD, indicating that OCD risk extends to genes regulated by *CHD8* (Table 2 and eFigure 2 in Supplement 1).

Given our findings with rare variation, we next explored whether *CHD8* pathway genes show an enrichment of common variation signal, identified in the most recent OCD genome-wide association study (GWAS) including 53,660 OCD cases and 2,044,417 controls of European ancestry.^69^ Using Multi-marker Analysis of GenoMic Annotation^51^ (MAGMA v1.10), single-nucleotide polymorphisms (SNPs) were mapped to 19,427 protein coding genes. We performed gene-set enrichment analysis using the six *CHD8* pathway gene lists^64,65^ and observed three lists derived from Cotney et al. 2015 reached Bonferroni-corrected significance (*P*<0.008) (Table 3). The gene-set analyses also implicated common variants in *CHD8* pathway genes in ASD, bipolar disorder, schizophrenia, and attention-deficit/hyperactivity disorder (ADHD) (eTable 8 in Supplement 2).

**Table 3.** MAGMA gene-set enrichment analysis using *CHD8* pathway gene lists and OCD GWAS. . NGENES (the number of genes overlapping between *CHD8* pathway genes and the GWAS), BETA (effect size) and standard error of BETA (SE), as well as MAGMA *P* value are shown for each list. Gene lists with *P* value < 0.05 are bolded. For additional results, see Supplemental Table 8. *CHD8*, chromodomain-helicase-DNA-binding protein 8; Cotney_brain, *CHD8* target gene list identified in human midfetal brain cells (Cotney et al., 2015); Cotney_hNSC, *CHD8* target gene list identified in human neural stem cells (Cotney et al., 2015); OCD, obsessive-compulsive disorder.

| VARIABLE | MAGMA |  |  |  |
| --- | --- | --- | --- | --- |
| | NGENES | BETA | SE | $P^*$ |
| ChIP-seq analysis in hNSC from Cotney et al., 2015 | 8,367 | 0.050 | 0.014 | **$1.85 \times 10^{-4}$** |
| ChIP-seq analysis in Brain from Cotney et al., 2015 | 2,490 | 0.066 | 0.020 | **$3.85 \times 10^{-4}$** |
| ChIP-seq analysis in hNSC&Brain from Cotney et al., 2015 | 2,306 | 0.071 | 0.020 | **$2.13 \times 10^{-4}$** |

*ZMYM2* emerged as an important gene in the shared genetics of OCD and CTD. *ZMYM2* encodes a transcription factor, and four studies have identified genes with altered expression following homozygous disruption of *ZMYM2* in stem cells.^70–73^ Using gene-set analysis of genes showing altered expression in two or more of these studies (n = 168), we found that 18% of *ZMYM2*-regulated genes are likely implicated in OCD (*P* = 0.048) (Table 2). Furthermore, MAGMA analyses suggest that common variation in *ZMYM2*-regulated genes may also contribute to ADHD risk (eTable 8 in Supplement 2). These findings highlight that, in addition to *CHD8*, *ZMYM2* and its downstream pathways may play a role in the genetic architecture of OCD and related neurodevelopmental disorders.

## Discussion

This study represents the largest analysis of rare coding variation in OCD, providing novel insights into the genetic architecture of this disorder. By assembling a dataset six times larger than previous sequencing studies, our work is the first to achieve sufficient power to identify a genome-wide significant OCD risk gene, confirming the role of *CHD8* in OCD susceptibility. We demonstrated that deleterious mutations in highly conserved genes play a significant role in OCD risk, with an estimated ∼470 autosomal genes implicated in its susceptibility. Genetic risk for OCD extends beyond SNVs, evidenced by the significant enrichment of rare genic CNVs identified in both our de novo and case-control analyses. While large CNVs have been implicated in OCD,^15^ this is the first study to demonstrate a significant contribution from small CNVs, particularly deletions affecting highly evolutionarily constrained genes. These findings suggest reduced gene dosage is critical for OCD etiology, complementing our analyses that implicate deleterious single-nucleotide variants in the disorder.

Four lines of evidence together provide compelling support for a role for *CHD8* and *CHD8* pathways genes in OCD. First, we identified *CHD8* as a significant OCD risk gene (q = 0.038), highlighting its crucial and wide-ranging role in psychiatric disorders. In addition, we observed that both rare and common variation in genes targeted by *CHD8* were associated with OCD risk. Finally, we leveraged RNAseq data from a parallel study of postmortem samples from individuals with OCD or Eating Disorders (ED) and found that there were significant differences in expression of *CHD8* targets in OCD and in ED^74^. These differentially expressed genes mapped to defined co-expression models in the caudate and in the dorsolateral prefrontal cortex that are involved in cellular energy production, providing a window into potential functional changes underlying OCD. This is the first time that multiple lines of evidence converge on a single risk pathway in OCD.

Our meta-analysis of OCD and CTD further identified *CHD8* and *CELSR3* as risk genes with q values less than 0.05 in the shared genetic architecture of these two disorders. Additionally, *WWC1* and *QRICH1* were identified with q values less than 0.1, supporting their potential role in overlapping pathways contributing to OCD and CTD risk.

Our gene-set analyses revealed biological pathways implicated in OCD and shared OCD-CTD genetic architecture, particularly transcriptional regulation, chromatin remodeling, and proteostasis. *CHD8,* a DNA-binding chromatin remodeling gene, regulates numerous downstream genes implicated in neurodevelopment, with both rare and common variation in these genes associated with OCD risk. Similarly, *ZMYM2* was implicated in both OCD and shared OCD-CTD genetic risk. *ZMYM2* is a zinc finger transcriptional regulator that appears to act as a transcriptional repressor; heterozygous protein truncating and missense variants in this gene are implicated in syndromic developmental disorders with a CNS phenotype.^71,75,76^

Proteostasis, the process of maintaining protein homeostasis, has been implicated in several neurodevelopmental disorders, including ASD and specific developmental delays such as Angelman syndrome which is caused by mutations in the E3 ubiquitin ligase gene *UBE3A,* a gene that regulates protein degredation.^76^ In the shared genetic architecture of OCD and CTD, proteostasis-related genes emerged as important contributors. *QRICH1* is involved in the transcriptional control of proteostasis and has been associated with developmental delay and autism, highlighting its pleiotropic effects.^77,78^ *HECTD4* is a E3 ubiquitin ligases that is relatively understudied, but E3 ligases are critically involved in neural development through proteostasis, and heterozygous and biallelic variants in *HECTD4* have been implicated in developmental delay.^80–82^

Together, our findings extend the relevance of proteostasis to OCD and with the shared genetic architecture of OCD and CTD, suggesting that disruptions in protein homeostasis may also play a broader role in psychiatric disorders, including schizophrenia. Ongoing studies of proteostasis as a therapeutic target in neurodegenerative disorders may provide valuable insights into its role in neurodevelopment and psychiatric illnesses. Disruptions in these cellular processes, including protein homeostasis and transcriptional regulation, may underlie the shared genetic architecture of OCD and related disorders. Further studies focusing on neuronal targets of *HECTD4, QRICH1*, and *ZMYM2* are needed to clarify their contributions to OCD pathophysiology and may provide insights into novel therapeutic strategies.

Our findings have important clinical implications, particularly for advancing precision medicine approaches in OCD and related disorders. The identification of specific risk genes, their biological processes, their relevant downstream targets, and their dosage effects provide a foundation for understanding the molecular mechanisms underlying OCD risk and for developing improved therapeutic strategies. Cross-disorder genetic analyses may further uncover shared pathways that could guide interventions for overlapping neuropsychiatric phenotypes, such as comorbid OCD, ASD, and CTD.

This study has several notable strengths. It represents the largest analysis of rare coding variation in OCD to date, leveraging data from 11 cohorts and over 44,000 individuals, including 6,071 individuals diagnosed with OCD. By examining both rare SNVs and CNVs, including small genic deletions, this study provides a comprehensive view of rare variation contributing to OCD risk. The study’s integration of pathway-level analyses proposes key molecular mechanisms underlying OCD and its shared genetic architecture with related neuropsychiatric disorders. Limitations of this study should also be acknowledged. First, while this is the largest analysis of rare coding variation in OCD to date, the sample size limits the detection of smaller effect-size genes or variants. Second, the focus on exonic regions excludes rare variants in non-coding regulatory regions, which may also play a significant role in OCD risk. Third, although significant enrichments in constrained genes were observed and we identified putative candidate genes, functional validation will be necessary to confirm their roles and elucidate their mechanisms in OCD pathophysiology. Finally, the generalizability of the findings may be limited by the predominantly European ancestry of the sample population, underscoring the broad for studies in more diverse populations to improve applicability.

In conclusion, this study provides new insights into the genetic architecture of OCD, identifying the enrichment of deleterious rare coding variants and structural variants in highly constrained genes. It is well-established that OCD shows high comorbidity with CTD and other pediatric onset psychiatric disorders such as ASD and DD. *CHD8*, in particular, emerges as a critical gene and pathway in OCD, and as a node linking OCD to broader neurodevelopmental and psychiatric conditions. It is somewhat less appreciated that OCD is associated with as much as a 10-fold increased risk for a comorbid schizophrenia or bipolar disorder diagnosis, and longitudinal analyses show that a first diagnosis of OCD is also highly associated with elevated risk for a subsequent schizophrenia, schizoaffective, or bipolar disorder diagnosis.^15,66–68^ Our results underscore the shared genetic architecture of these neuropsychiatric disorders, providing a window into the complexity and pleiotropy of the genetic underpinnings of these conditions.

## Ethics statement

This study was approved by the Regional Ethical Review Board in Stockholm (Dnr 2013/1849–31/2) and the Institutional Review Board (IRB) at the Icahn School of Medicine at Mount Sinai, New York, NY. All participants provided informed consent.

## Acknowledgements

We want to thank all participants and families that were a part of the study cohorts used in this research.

This study was supported by NIH grants MH124679 (DEG), MH124675 (JJC), and MH129724 (JDB), and by an IOCDF Innovator Award to DEG. The IOCDF Innovator Award was funded by the Walder Family Charitable Fund.

The All of Us Research Program is supported by the National Institutes of Health, Office of the Director: Regional Medical Centers: 1 OT2 OD026549; 1 OT2 OD026554; 1 OT2 OD026557; 1 OT2 OD026556; 1 OT2 OD026550; 1 OT2 OD 026552; 1 OT2 OD026553; 1 OT2 OD026548; 1 OT2 OD026551; 1 OT2 OD026555; IAA #: AOD 16037; Federally Qualified Health Centers: HHSN 263201600085U; Data and Research Center: 5 U2C OD023196; Biobank: 1 U24 OD023121; The Participant Center: U24 OD023176; Participant Technology Systems Center: 1 U24 OD023163; Communications and Engagement: 3 OT2 OD023205; 3 OT2 OD023206; and Community Partners: 1 OT2 OD025277; 3 OT2 OD025315; 1 OT2 OD025337; 1 OT2 OD025276. In addition, the All of Us Research Program would not be possible without the partnership of its participants.

The BioMe Biobank is supported by The Charles Bronfman Institute for Personalized Medicine at Mount Sinai.

This research has been conducted using the UK Biobank Resource under application number 84208.

This work was supported in part through the computational and data resources and staff expertise provided by Scientific Computing and Data at the Icahn School of Medicine at Mount Sinai and supported by the Clinical and Translational Science Awards (CTSA) grant UL1TR004419 from the National Center for Advancing Translational Sciences. Research reported in this publication was also supported by the Office of Research Infrastructure of the National Institutes of Health under award number S10OD026880 and S10OD030463. The content is solely the responsibility of the authors and does not necessarily represent the official views of the National Institutes of Health.

## Data and code availability

All rare variant findings used in reported analyses (including SNV and gCNV calls) are included in Supplemental Tables. Researchers may request access by contacting Dr. Joseph D. Buxbaum. All requests will be reviewed by the Mount Sinai Institutional Data Access Committee to ensure compliance with participant consent and IRB protocols.

