## Supplement_1 for "Rare coding variation in OCD implicates shared genes with other psychiatric disorders"

**Supplemental Online Content**

**Consortium authors**

***Epidemiology and Genetics of OCD and chronic tic disorders in Sweden (EGOS)***

Behrang	Mahjani

Karin	Dellenvall

Anna-Carin	Säll	Grahnat

Gun	Karlsson

Aki	Tuuliainen

Christina	G	Mahjani

Lambertus	Klei

Silvia	De	De Rubeis

Marina	Natividad	Avila

Laura	Sloofman

Jacob	Gluckman

Abraham	Reichenberg

Bernie	Devlin

Christina	M	Hultman

Joseph	D	Buxbaum

Sven	Sandin

Dorothy	E	Grice

***Nordic OCD and Related Disorders Consortium (NORDiC)***

Julia	Bäckman

Long	Long	Chen

James	J.	Crowley

Elles	de Schipper

Diana	R.	Djurfeldt

Jan	Haavik

Kristen	Hagen

Matthew	W.	Halvorsen

Bjarne	Hansen

Kira	D.	Höffler

Fredrik	Johansson

Anna	K.	Kähler

Elinor	K.	Karlsson

Gerd	Kvale

Paul	Lichtenstein

Kerstin	Lindblad-Toh

Manuel	Mattheisen

David	Mataix-Cols

Christian	Rück

Thorstein	Olsen	Eide

Nora	I.	Strom

John	Wallert

**eMethods**

**Joint genotyping**

**EGOS and LifeGene control cohorts**

The previously unpublished 2,115 samples from EGOS cohort were sequenced on Illumina HiSeq sequencers using the Twist Human Comprehensive Exome Kit following standard protocols (paired-end, 2 × 150 bp reads). Exome sequencing of 4,765 Swedish LifeGene controls was performed at the Broad Institute of MIT and Harvard. We removed the adapter sequence from WES data of EGOS and LifeGene control cohorts using Trimmomatic v.0.36.^1^ The sequencing reads in trimmed fastq files of EGOS and LifeGene control cohorts were aligned to the same human genome build 38 (GRCh38) using bwa v.0.7.15, aggregated to bam files.^2^ We used Picard v.2.22.3 (http://broadinstitute.github.io/picard/) to sort chromosome coordinates and mark duplicates for bam files. To get more accurate base quality scores in bam files, base quality score recalibration (BQSR) in Genome Analysis Toolkit (GATK) was applied following the GATK Best Practices.^3^ SNVs and indels were called using local realignment by HaplotypeCaller in GATK in gVCF mode based on individual bam file. We combined whole gVCFs from the EGOS and LifeGene control cohorts and performed joint genotyping using GenotypeGVCFs of GATK.

**Mount Sinai BioMe Biobank, Yale, and Columbia/Johns Hopkins cohorts**

For the Columbia/Johns Hopkins cohort raw fastq files were trimmed and aligned to GRCh38 human reference genome using Trimmomatic and bwa, as described for the EGOS and LifeGene cohorts. For the Yale and Mount Sinai BioMe Biobank cohorts, we used mapped bam files. Specifically, the bam files of the Yale cohort, which originally were aligned to GRCh37 reference, were converted to GRCh38 using CrossMap v.0.6.3.^4^ For the Columbia/Johns Hopkins, Yale, and Mount Sinai BioMe Biobank cohorts, we carried out the subsequent processes of sorting, marking duplicates, BQSR, making gVCFs, joint-genotyping, and estimation of variant call accuracy using Picard and GATK , following the methodologies previously mentioned.

**Quality control**

**Multiallelic Site Treatment and Annotation:** We first addressed multiallelic sites in the VCF file, splitting them into bi-allelics by the LeftAlignAndTrimVariants function in GATK. These variants were then annotated using the Variant Effect Predictor (VEP) to predict the potential functional effects ^5^.The tool predicts how the variant might alter the amino acid sequence, affect splicing, introduce stop codons, or affect regulatory elements, among other possibilities.

**Data Transformation and Pedigree Verification:** We imported the VCF file into Hail 0.2 (https://hail.is) and converted it to a Matrix Table for quality control (QC) process. We calculated relatedness between each pair of samples using an identity by descent (IBD) function of Hail to verify reported pedigrees of family data and check for duplicate samples within and across datasets. Sex of each sample was imputed using impute_sex() function. The relatedness and sex information was used as input data for Rapid Reconstruction of Pedigrees from Genome-wide Estimates of Identity by Descent (PRIMUS) to infer the pedigree.^6^ Obvious errors in reporting were fixed (e.g., swapped mother/father or parent/child labels in the same family). Trios with a discrepancy that could not be resolved (trios without a child in the dataset, individuals showing pi-hat > 0.2 with all other individuals in the IBD test, and individuals with mis-matched information between imputed- and reported-sex) were dropped. For variant QC, we removed variants in the low-complexity regions or those not passing VQSR.

**Genotype and Sample Quality Control:** Comprehensive QC was undertaken for both genotypes and samples. For genotype QC, we removed genotype calls using several filters: genotype calls with a depth below 10 or above 1,000; genotype calls in the Y chromosome from female samples; homozygous reference calls with less than 90% of the read depth supporting the reference allele or with a genotype quality (GQ) < 25; heterozygous calls with the phred-scaled likelihood of the call being homozygous reference (PL[HomRef]) < 25, with call rate (the read depth supporting either the reference or alternate allele) < 90%, with the allele balance (number of mapped reads supporting the alternate allele divided by the read depth) < 25, with a probability of the allele balance based on binomial distribution centered on 0.5 less than 1×10^9^ or in the X or Y chromosome (excluding pseudoautosomal regions) of male samples; homozygous alternate calls with less than 90% of the read depth supporting the alternate allele or with a PL[HomRef] < 25. We removed samples with high proportion of contaminated reads (FREEMIX > 7.5%) estimated by verifyBamID ^7^, samples with a proportion of chimeric reads > 7.5% estimated by CollectAlignmentSummaryMetrics function in Picard v.2.22.3 (http://broadinstitute.github.io/picard/), samples with a call rate greater than 3 standard deviations below the mean of each cohort, duplicated samples, and samples showing close genetic relatedness to other samples in the case-control cohorts (kinship value > 0.1 in KING).^8^ In the family-based data, probands whose one or both parents had been removed were reclassified as cases in the case-control data and the remaining parent (if any) was removed, and if one parent was filtered and no proband remained, the other parent was removed. For the final variant QC, we removed variants with a call rate < 10%, or with a Hardy-Weinberg equilibrium (HWE) *P* value < 10^-12^. After QC, the VCF of EGOS, LifeGene control, Mount Sinai BioMe Biobank, Yale, and Columbia/Johns Hopkins cohorts contained a total of 9,741 samples including 725 OCD trios, 40 OCD quartets, 2,110 OCD cases and 5,296 controls. A summary of the quality control and data cleaning procedures of the joint genotyping data of EGOS and LifeGene control cohorts, and Mount Sinai BioMe Biobank, Yale, and Columbia/Johns Hopkins cohorts can be found in Method Figure 1a and b, respectively.

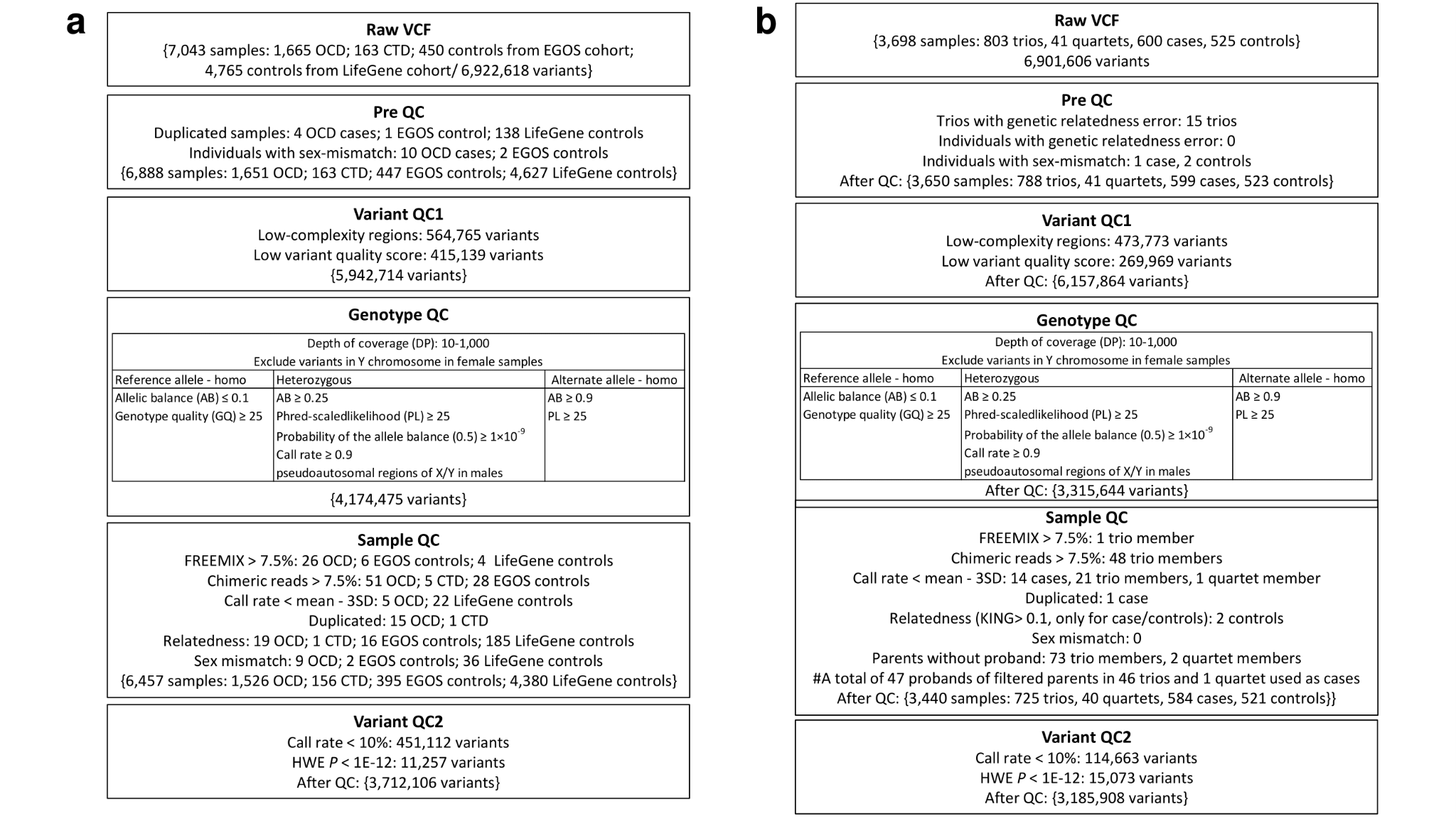

**Method Figure 1. Flow chart of quality control (QC).** **a)** Details of QC of joint genotyped WES data using EGOS and LifeGene control cohorts. **b)** Details of QC of joint genotyped WES data using Mount Sinai BioMe Biobank, Yale, and Columbia/Johns Hopkins cohorts.

**De novo variants calling**

We removed 14 OCD trios in Yale that overlapped with samples in the Wang et al., 2018 cohort for both de novo and inherited variants calling. A total of 791 trios (711 trios + 40 quartets) were used for the further analysis. The de_novo() function in Hail v0.2.60 (https://hail.is) was used to identify de novo variants from the QC-ed joint-genotyped data (removing variants with a GQ < 25). The population allele frequency of the non-neuro subset from the gnomAD database ^9,10^ was used as the input priors of the de_novo() function. We also used “max_parent_ab = 0.03”, “min_child_ab = 0.3”, and “min_dp_ratio = 0.3” as parameters of the de_novo() function to exclude de novo candidates if the allele balance in a parent is above 3%, if the allele balance of proband is lower than 30%, or if the ratio of read depth between the proband and parents is lower than 30%. This process yielded 7,009 putative de novo variants from a total of 791 probands.

**Quality Control on De Novo Variants:** From the initial 7,009 variants, we kept 2,602 variants with “HIGH” or “MEDIUM” confidence indicated by the calling algorithm (the “MEDIUM” confidence calls are limited to a singleton). We further removed 188 and 145 variants with allele frequency > 0.1% in the QC-ed joint-genotyped data and non-neuro subset from the gnomAD database, respectively. Then, 3 variants were excluded as they appeared more than twice, and additional 205 variants were removed to retain one variant with the most severe effect in the same gene for each sample. Finally, 170 variants identified from 16 children with >7 de novo variants were removed for the consistency with the cutoff of de novo count per sample at least >7 in the 5,456 unaffected siblings.^11^ After QC, a total of 1,891 high confidence de novo variants of 791 trios remained from Yale and Columbia/Johns Hopkins cohorts (eTable 2 in Supplement 2).

**Incorporation of External Data:** In addition to our data, we incorporated 54 de novo variants from 53 OCD trios in Lin et al., 2022, 242 de novo variants from 306 OCD trios in Wang et al., 2018, 68 de novo variants from 52 OCD trios in SPARK Consortium, and 9,034 de novo variants from 5,492 unaffected sibling trios in Fu et al., 2022 in this study (eTable 2 in Supplement 2).

**Rare inherited variants calling**

We used the transmission_disequilibrium_test() function in Hail to identify rare inherited variants. This function measures how often the alternate allele is transmitted versus untransmitted from heterozygous parents to an affected child. Our analysis drew upon the 791 trios that met the quality criteria during the de novo variant QC stage.

**Genotype and Variant Filtering:** Genotype calls were filtered using thresholds set at GQ > 25 for all genotypes and allele balance > 0.3 specifically for heterozygous genotypes. For filtering variants, we kept variants with a call rate > 90% and single nucleotide variants with a VQSLOD (variant quality score log odds) ≥ -1.60 in Yale and 1.04 in Columbia/Johns Hopkins cohort. The VQSLOD threshold for SNVs was determined by identifying the threshold at which synonymous variants with an allele count of 1 among parents in the dataset were transmitted to the child 50% of the time.^12,13^ To keep PTVs with a high level of confidence, we used the LOFTEE (Loss-Of-Function Transcript Effect Estimator) plugin, a tool designed to predict the functional effects of genetic variants that potentially cause loss-of-function in genes. This plugin is used in conjunction with the VEP tool. The LOFTEE plugin for VEP mandated that PTVs should have a "HC" designation and must not possess any LOFTEE flags other than "SINGLE_EXON."

The transmitted or untransmitted variants were required to have an allele count of ≤5 in the variant list for our entire case-control cohort (a total of 24,115 individuals), and in the non-psychiatric subset of the gnomAD database ^14^.

**Rare case-control variants calling**

**EGOS, LifeGene control, BioMe Biobank, and Columbia/Johns Hopkins cohorts**

To ascertain the ancestry of EGOS and LifeGene cohorts comprising 1,526 OCD cases and 4,775 controls, we conducted principal components analysis (PCA) using “smartpca” of EIGENSOFT.^15^ For this analysis, we kept variants that had undergone LD pruning, had a call rate > 99%, and a minor allele frequency > 0.001 from the QC-ed joint-genotyping data, and then merged them with 1000 Genomes reference data. A total of 201 outliers (30 cases and 171 controls) in PCA were removed automatically by “smartpca”. To predict ancestries of EGOS and LifeGene samples, we utilized a random forest classifier.^16^ Then, using the remaining 1,496 cases and 4,604 controls, we performed 1:2 case:control matching based on sex and the first five principal components. The matching was performed using the function “match_on” in the R package of “optmatch” ^17^ (Method Figure 2a). We kept 1,496 OCD cases and 2,992 controls from the QC-ed joint-genotyping data for the further analyses. For the case-control coverage harmonization, we kept variants in high coverage, defined as call rate ≥90% and depth of coverage ≥10 in both cohorts. To check the coverage harmonization, we checked quantile-quantile plots of synonymous and protein-truncating variants (Method Figure 3a and b). We employed a permutation-based assessment using the QQperm R package (https://cran.r-project.org/web/packages/QQperm/index.html). This package utilizes a gene-by-sample collapsing analysis matrix, permuting case–control labels to generate expected chi-square statistics. This process involved 10,000 permutations in which we permuted case-control labels within each distinct case-control group, computed gene-based test statistics for the permuted datasets using the Cochran–Mantel–Haenszel test as previously described, and obtained the median chi-square statistic from the set of tests. We confirmed there was no genomic inflation (λ ≈ 1) in both synonymous variants and protein-truncating variants (Method Figure 3a and b).

In the Mount Sinai BioMe Biobank cohort, we confirmed that 105 cases and 525 controls were well-matched based on sex and ancestry (Method Figure 2b). We applied certain filters to genotype calls, including a GQ threshold of >25 for all genotypes and an allele balance threshold of >0.3 for heterozygous genotypes. Variants were required to have a call rate of ≥ 90% and pass VQSR. For filtering rare variants, the variants had to have an allele count of ≤5 in the variant list of the entire case-control cohort as well as in the non-psychiatric subset of the gnomAD database.^14^ For matched case-control data of Columbia/Johns Hopkins cohort, we used rare variant counts from the previous paper ^18^.

**All of Us Research Program**

All of Us Research Program (All of Us) is a prospective cohort study targeting the recruitment of a diverse group of at least 1 million individuals in the United States to accelerate biomedical research and improve health.^19^ On April 20, 2023, All of Us released short read whole genome sequencing (WGS) data of 245,394 samples (v7 data release) within the Researcher Workbench for use by researchers registered for Controlled Tier access.

Among these 245,394 samples, we identified 1,609 OCD cases using either the ICD10 code of F42 or ICD9 code of 300.3, and 156,810 controls without any mental disorders. Then, 1,609 OCD cases were matched to 8,045 controls based on sex and ancestry (PC1-5) using a R package of “optmatch” to yield 1:5 case:control matches (Method Figure 2c). From these samples, we extracted information from the Hail multi MatrixTable for the exome short read WGS joint callset (gs://fc-aou-datasets-controlled/v7/wgs/short_read/snpindel/exome/splitMT/hail.mt).

For functional annotations of variants in All of Us data, we used VEP as described above.^5^ We Adhered to the guidelines set by the All of Us for sample selection and variant calls (https://support.researchallofus.org/hc/en-us/articles/4617899955092-All-of-Us-Beta-Release-Genomic-Quality-Report). We removed 16 erroneous samples and variants with low quality.

In our own sample QC process, we removed samples with a call rate greater than 3 standard deviations below the mean, duplicated samples, and samples showing close genetic relatedness to other samples (kinship value > 0.1 in KING). After the sample QC, 1,595 OCD cases and 7,944 controls were left.

For the genotype QC, we excluded homozygous reference calls with less than 90% of the read depth supporting the reference allele or with GQ < 25; heterozygous calls with the allele balance < 30, with a GQ < 25, or with a probability of the allele balance based on binomial distribution centered on 0.5 less than 1×10^9^; homozygous alternate calls with less than 90% of the read depth supporting the alternate allele or with GQ < 25. Variants were required to have a call rate of ≥ 90% and a HWE *P* value > 10^-12^. For filtering rare variants, the variants had to have an allele count of ≤ 5 in the variant list of the entire case-control cohort as well as the non-psychiatric subset of the gnomAD database.

**UK Biobank**

UK Biobank (UKB) is a comprehensive prospective study with over 500,000 participants with extensive and readily accessible phenotypic and genetic data.^20^ From 502,387 participants enrolled in the UKB workspace, we found 1,253 OCD cases with an ICD10 code of F42 or ICD9 code of 300.3, and 282,740 controls without any mental disorders. Based on sex and the first five principal components, the 1,253 OCD cases were matched to 6,265 controls using an R package of “optmatch” (Method Figure 2d). (file:////mnt/project/Bulk/Exome sequences/Population level exome OQFE variants, pVCF format - final release/).

For the sample QC, we removed samples with a call rate greater than 3 standard deviations below the mean, duplicated samples, and samples showing close genetic relatedness to other samples (kinship value > 0.1 in KING). After the sample QC, 1,212 OCD cases and 6,028 controls were left.

For genotype QC, we removed genotype calls using the same hard filters applied in the All of Us data. We generated functional annotations of high quality variants using VEP.^5^ For filtering rare variants, the variants had to have an allele count of ≤ 5 in the variant list of the entire case-control cohort as well as the non-psychiatric subset of the gnomAD database.

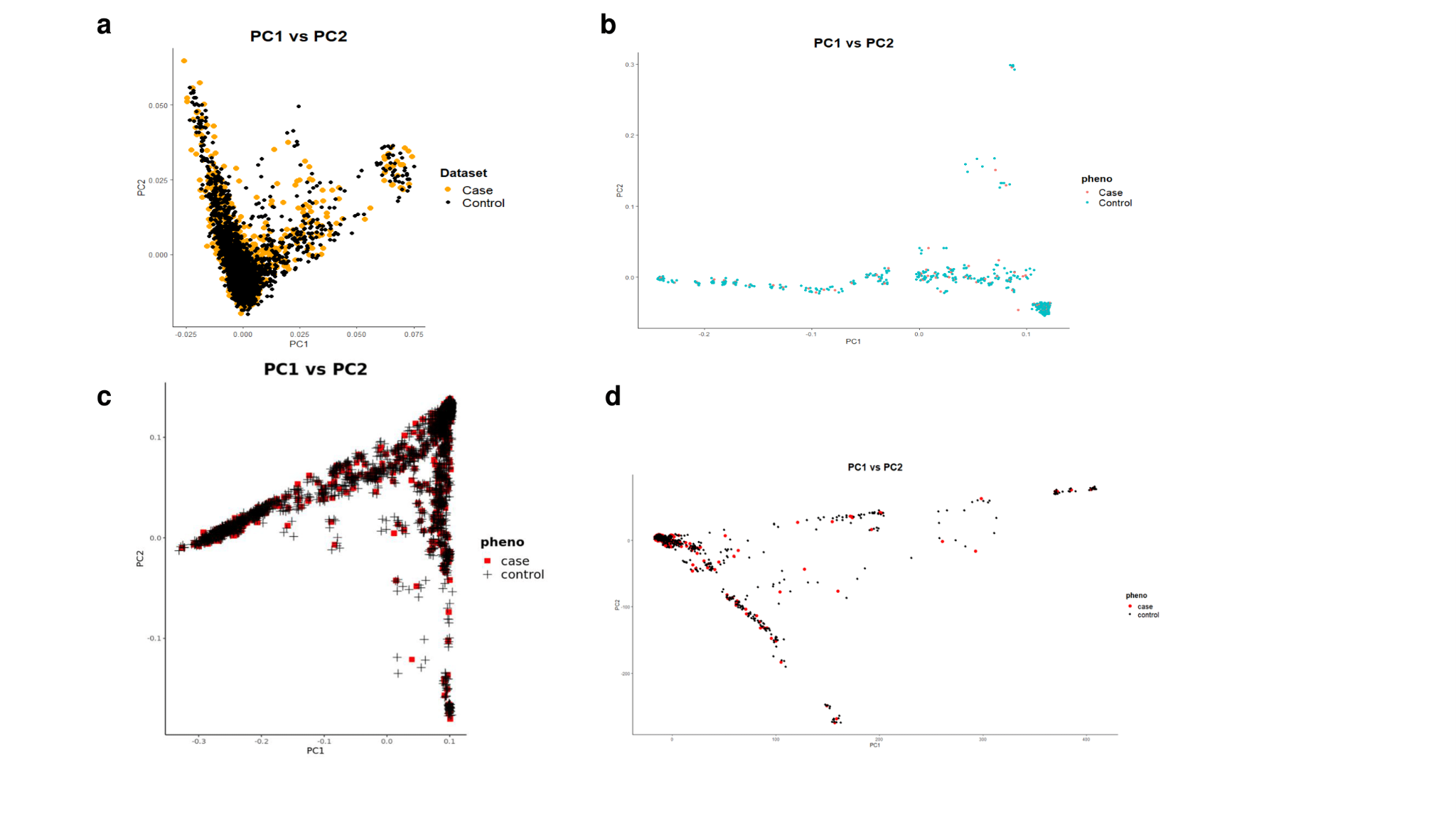

**Method Figure 2. Principal components analysis plots after case-control matching.** **a)** 1,496 EGOS cases and 2,992 LifeGene controls, **b)** 105 cases and 521 controls in the Mount Sinai BioMe Biobank cohort, **c)** 1,595 cases and 7,944 controls in the AllofUs cohort, and **d)** 1,212 cases and 6,028 controls in the UK Biobank cohort.

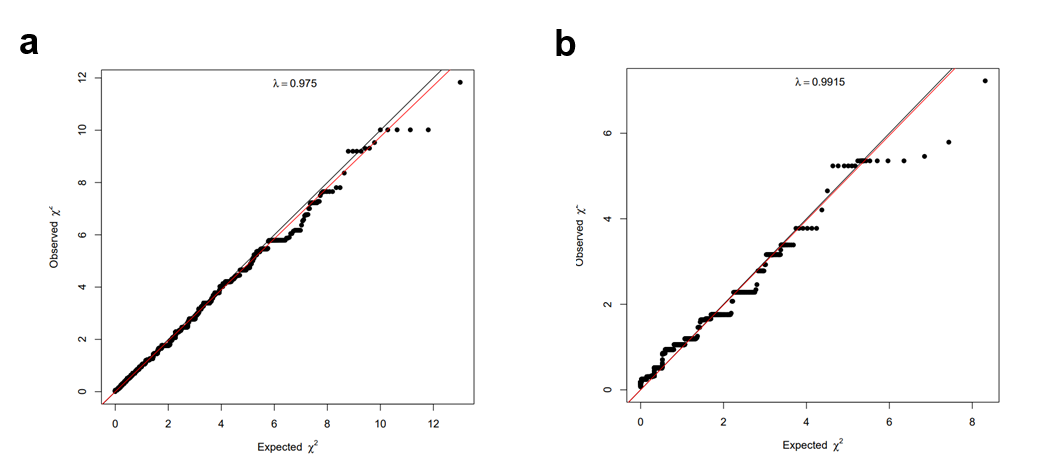

**Method Figure 3. Quantile-quantile (QQ) plot from a gene-based collapsing using EGOS and LifeGene dataset.** Gene-based test statistics under a) synonymous variant, and b) protein-truncating variant model.

**CNV analysis**

From a combined total of QC-ed 7,247 samples including 791 OCD trios from Yale and Columbia/Johns Hopkins cohorts, 1,601 cases and 3,513 controls from EGOS, LifeGene control, and BioME BioBank Program cohorts, the GATK-gCNV method was employed for detecting germline CNVs.^21^

The initial step involved compressing raw sequencing files into read counts across annotated exons, which were then utilized as input data. Subsequently, an approach based on principal component analysis was utilized to differentiate variations stemming from distinct capture kits (Method Figure 4a-c). This was followed by a hybrid approach that combined density and distance-based clustering. This clustering method was used to curate batches of samples that could be processed in parallel. Cohort mode of GATK-gCNV was performed using 200 samples in every cluster determined by PCA. CNVs in the same cluster with a minimum of 80% reciprocal overlap were determined as the same location.

For quality control of CNVs, we kept variants with frequency less than 1% and that spanned more than two captured exons. For homozygous deletions, the QS threshold is set to the smaller value between 400 and ten times the number of intervals; for heterozygous deletions, the QS threshold is set to the smaller value between 100 and ten times the number of intervals; and for duplications, the QS threshold is set to the smaller value between 50 and four times the number of intervals.

For sample-level quality control, samples were kept if their number of raw, autosomal CNV calls detected by GATK-gCNV did not exceed 200 and if the number of calls with QS≥20 did not exceed 35. After QC, a total of 7,098 samples remained, consisting of 676 OCD trios, 1,598 cases and 3,472 controls. We considered a gene to be impacted by deletion if at least 10% of the non-redundant exons of the gene were overlapped by the deletion. For duplication, we considered a gene to be impacted if at least 75% of the non-redundant exons were overlapped by the duplication. We also annotated CNVs against a list of 79 curated genomic disorder (GD) loci.^11^ A CNV call was considered a genomic disorder CNV if it shared at least 50% reciprocal overlap with the annotated GD. To identify de novo CNVs, we first analyzed the CNVs of all three family members. We filtered the CNVs in the offspring to keep only high-quality CNVs that were not present in either parent or were only partially present (less than 30% of genomic coordinates). Then, we kept high quality de novo CNVs with QS ≥ 220 regardless of the size. A total of 20 CNVs in 676 OCD trios were determined as high quality de novo CNVs and, for de novo CNVs of unaffected sibling trios, we used 72 de novo CNVs in 5,098 siblings (eTable 3 in Supplement 2).^11^ Conversely, high-quality CNVs in the OCD probands that overlapped with unfiltered CNVs in the parents were considered inherited CNVs if more than 30% of the captured intervals overlapped.

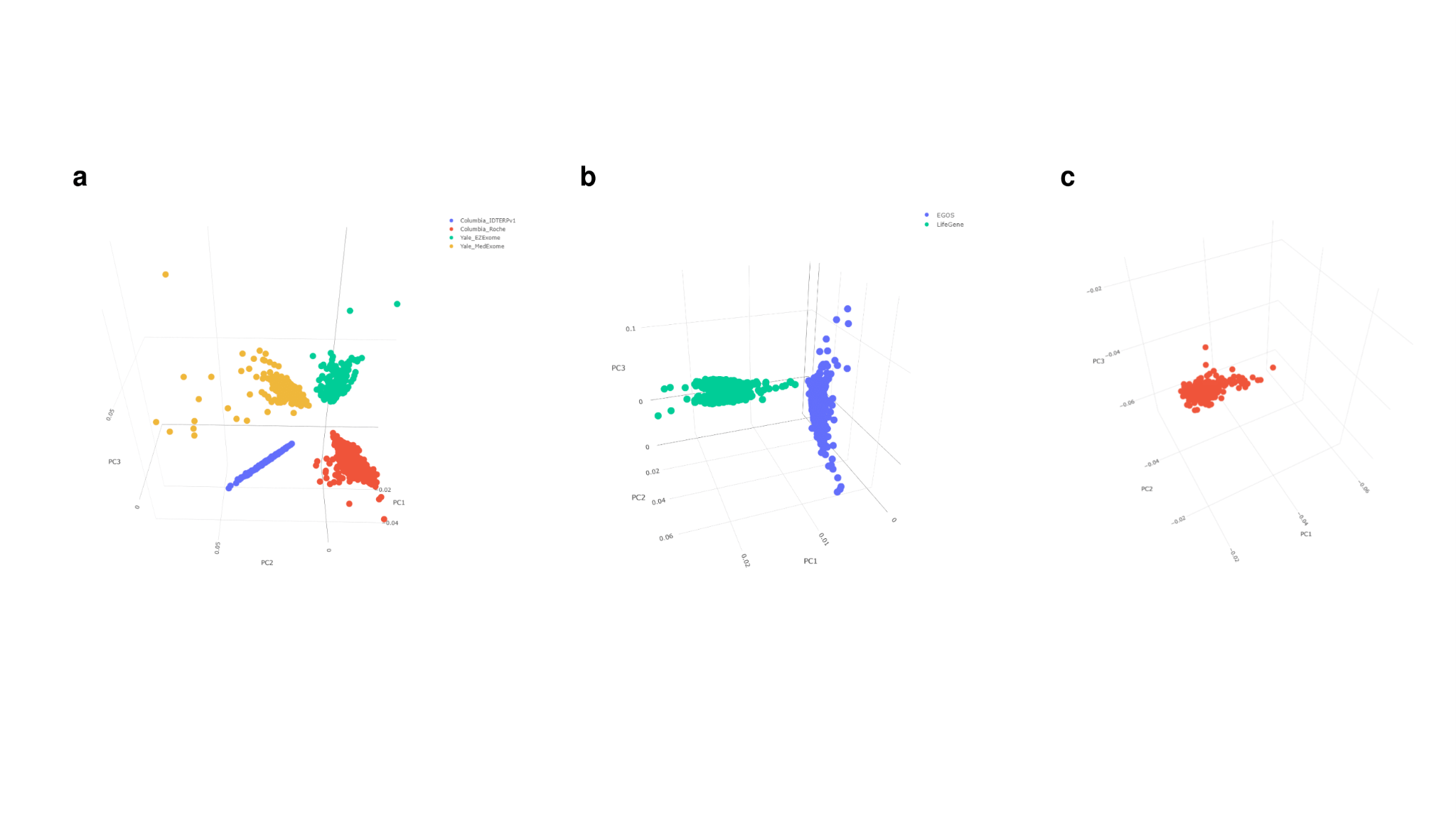

**Method Figure 4. Principal components analysis plots of read counts.** **a)** Trio WES data from Yale and Columbia/Johns Hopkins cohorts, **b)** Case-control WES data from EGOS and LifeGene control cohorts, **c)** Case-control WES data from BioME BioBank Program cohort.

**Transmission and De Novo Association Test (TADA)**

TADA is a Bayesian model that effectively combines de novo, transmitted/untransmitted, and case-control variants.^22^ Within this framework, TADA employs a Bayesian approach to generate gene-level indicators of association evidence, which can subsequently be converted into a measure known as the false discovery rate (FDR). In this current analysis, we used the extended TADA to integrate (1) rare coding variants, (2) rare CNVs, and (3) LOEUF^10^ scores.

In essence, TADA considers a particular gene and type of genetic variant. For rare genetic variants, we classified frameshift, stop gained, splice donor, splice acceptor and transcript ablation variants into protein-truncating variants (PTVs) based on the results of VEP.^5^ Missense variants were grouped by the MPC score: MisB variants with MPC ≥ 2, MisA variants with 2 > MPC ≥ 1, and Mis variants with 1 > MPC ≥ 0.^23^

To assemble a variant count table for TADA input, the number of rare de novo, transmitted/untransmitted, case-control variants, and CNVs that passed through QC filters were tallied for each gene (eTable 5 in Supplement 2). This variant count table encompassed variant classification (PTV, MisB, MisA, and synonymous variants), mutation rate per gene, and LOEUF score per gene.

TADA calculates a Bayesian Factor (BF) to quantify the statistical strength of evidence for gene-level association. The calculation of BF is based on various inputs, including the count of variant events, mutation rate, sample size, proportion of risk genes and a prior assessment of variant risk (gamma) within each gene. To estimate the proportion of risk genes for OCD, we used equation (29) and (33) from the supplementary method of Fu et al., 2022.^11^ For each gene, gamma PTV is calculated as "the relative risk of PTVs for each inheritance type" divided by "the estimated fraction of genes that substantially influence OCD risk", smoothed over LOEUF score. Using a sliding window of 20% of the genes, ordered by LOEUF, we computed a rolling-average gamma PTV for the fraction of risk genes: the relative enrichment of PTVs in probands compared to unaffected siblings for the de novo variants, the relative enrichment of transmitted PTVs compared to untransmitted PTVs for the inherited variants, the relative enrichment of PTVs in cases compared to controls for the case-control variants (eTable 5 in Supplement 2). To estimate the gamma for MisA and MisB variants for each gene, we first calculated the enrichment ratio of each variant class like gamma PTV above. We then subtracted 1 from the enrichment ratio, divided it by the estimated proportion of risk genes, and added 1, following the approach in Satterstrom et al., 2020 (eTable 5 in Supplement 2).^24^ To estimate the gamma for CNV, we added the sum of the gamma PTVs in constrained genes that are affected by that CNV. When we calculated the BF of deletions and duplications, we excluded NAHR-mediated or genomic disorder CNVs, and focused on modeling smaller CNVs that affect ≤ 8 constrained genes with LOEUF < 0.6.

To calculate the total gene-level BF, we multiply the BFs of the five variant classes: PTV, MisA, MisB, deletion, and duplication (all BFs of transmitted/untransmitted and MisA of case-control variants were excluded due to small effects). To ensure that evidence for association from one variant type is not weakened by evidence against association from another variant type, we set a minimum BF of 1 for each variant. To prevent genes from being nominated based on CNV evidence alone, we set the BFs for deletion and duplication to 1 if the total BF evidence for PTV, MisA, and MisB does not exceed 5. Finally, we converted BFs to posterior probabilities, which are then used to compute a q value to generate a list of potential candidate genes. Genes with an FDR less than 0.1 were classified as " high-confidence" risk genes, whereas those with an FDR below 0.3 were designated as "potential" risk genes, thresholds applied in rare variant studies.^18,24–26^

**Meta-analysis of OCD and CTD WES data**

For the meta-analysis, we calculated TADA parameters and BFs for each variant class for OCD and CTD separately, and then multiplied total BFs to identify common and unique risk genes between OCD and CTD (Method Figure 5). For CTD, we utilized rare coding variant counts from 523 trios across 3 existing WES studies (eTable 1 in Supplement 2).^27–29^ We created a count table, capturing the number of de novo mutations derived from the trios for every gene. This table encompassed variants classifications, (PTV, MisB, and MisA), mutation rate per gene, and LOEUF score per gene (eTable 6 in Supplement 2). We could not calculate the proportion of CTD risk genes, since no genes were identified with more than one de novo PTV event in our CTD WES data. Instead, we used the proportion of CTD risk genes at 0.04129527, as determined in the previously published CTD WES study by Wang et al., 2018.^27^ Using the variant count table, we calculate the CTD gamma value for de novo within each variant class per gene (eTable 6 in Supplement 2). Then, we calculated gene-level BFs for CTD in each variant class and merge them with BFs of OCD.

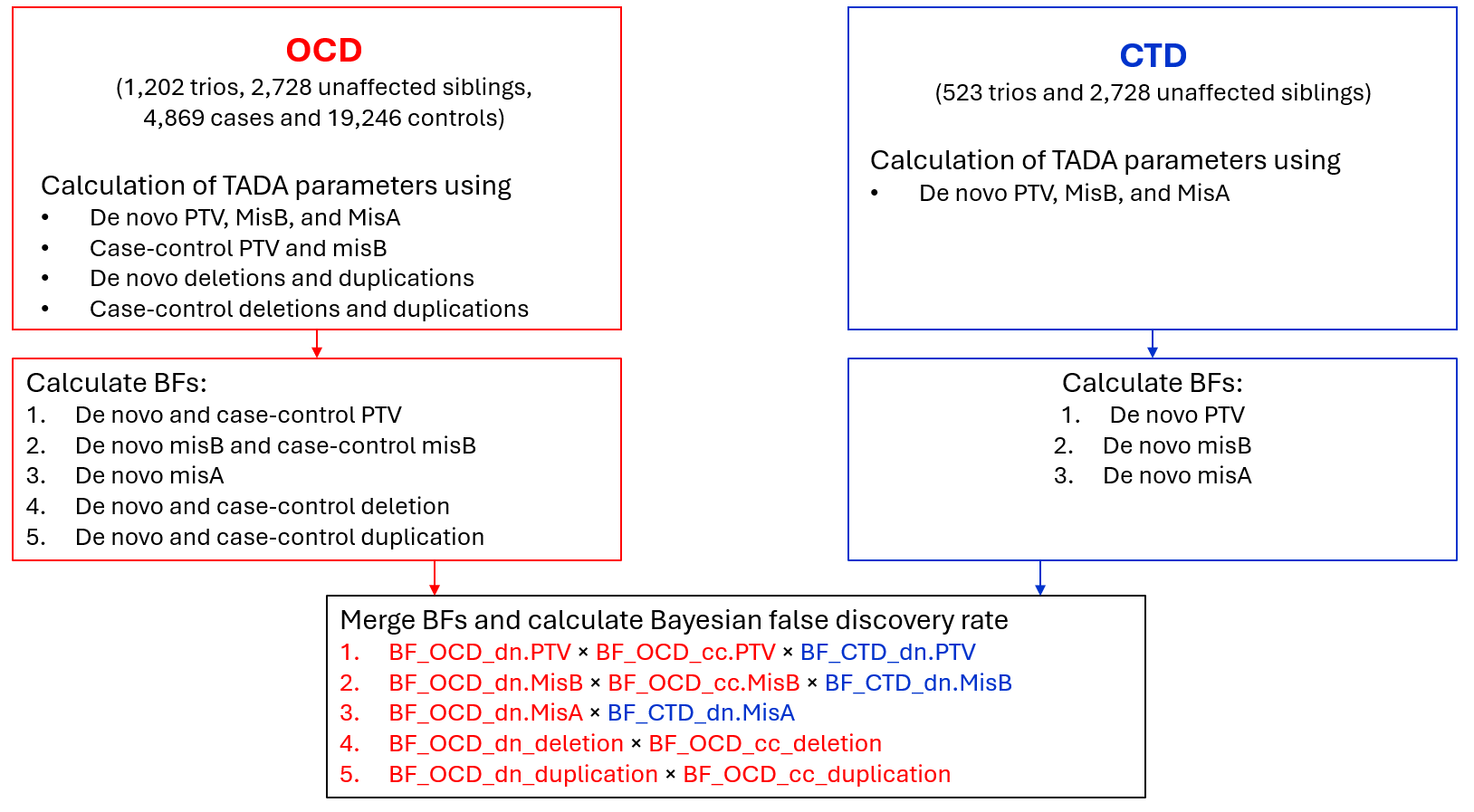

**Method Figure 5. Flow chart of meta-analysis using rare variant counts of OCD and CTD WES data.** TADA parameters and Bayes factors (BFs) for OCD and CTD were calculated separately by variant class, then combined to identify both shared and distinct risk genes. For CTD, data from 523 trios across three WES studies avoiding overlap with OCD study samples. De novo variant counts of 5,456 unaffected siblings from Fu et al., 2022 paper was divided into two sets through random selection, and each set was used separately for studies on OCD and CTD.

**Enrichment analysis**

To assess the overlap between OCD risk genes and the following gene lists: 1) CHD8 target genes; 2) ZMYM2 target genes; 3) autism spectrum disorder (ASD); 4) developmental delay (DD); 5) bipolar disorder; and 6) schizophrenia, we performed an enrichment analysis. We utilized four established CHD8 target gene lists, overlapped with genes in our OCD TADA result, from two independent studies: Cotney_brain (2,512 genes identified in human midfetal brain cells); Cotney_hNSC (8,451 genes identified in human neural stem cells); Wang_Neuron (3,145 genes identified in human early differentiating neurons); Wang_NPC (1,193 genes identified in human neural progenitor cells).^30,31^ For ZMYM2 target genes, we used 1,297 genes in the DEG analysis from Owen et al., 2023, 165 genes in the DEG analysis from Graham-paquin et al., 2023, 1,072 genes in the DEG analysis from Lezmi et al., 2020, and 544 genes in the DEG analysis from Tsusaka et al., 2020.^32–35^ We extracted risk genes of ASD and DD with FDR ≤ 0.1 in the TADA result from the most current ASD WES paper.^11^ Additionally, we selected the top 200 genes based on p-values for bipolar disorder and schizophrenia from existing studies.^14,36^ The selection of the top 200 genes aims to prioritize candidates that have demonstrated some degree of association or biological relevance in prior studies, even if they do not meet stringent statistical significance thresholds individually. While individual genes may lack strong statistical significance, they can still provide valuable insights when analyzed collectively, particularly if they are part of related pathways or biological processes. *P* values of our OCD TADA result were matched to the gene lists above. We estimated the proportion of true null hypotheses (π0) using the “propTrueNull” function in the limma package in R, denoted as from a set of *P* values obtained from multiple statistical tests. We used a method of “convest” in the propTrueNull function. Using π0, we calculated the proportion of alternative hypotheses (π1 = 1 – π0). The π1 larger than 0 indicates that there is a significant portion of hypotheses that are likely true alternatives, suggesting that there are non-random associations between OCD risk genes and the target gene lists. To assess the statistical significance of the overlap, a permutation-based method involving 1,000 random permutations was utilized and an empirical *P* value was calculated.

**Gene-set analysis**

We utilized MAGMA (v1.10) to perform gene-set analysis using GWAS summary statistics of OCD, ASD, schizophrenia (SCZ), bipolar disorder (BD) and attention-deficit/hyperactivity disorder (ADHD).^37–42^ Single nucleotide polymorphisms (SNPs) were mapped to genes using a window of 50 kb upstream and downstream of gene boundaries defined by NCBI genome assembly (GRCh37). Gene *P* value was computed by aggregating SNP *P* values within each gene, accounting for linkage disequilibrium estimated using the 1000 Genomes Project reference panel (European population). For gene sets, we used CHD8 or ZMYM2 target genes defined in the enrichment analysis above.^30–35^ Each gene set’s association with the trait was tested by comparing the mean of gene *P* values of the set against randomly sampled gene sets of the same size.

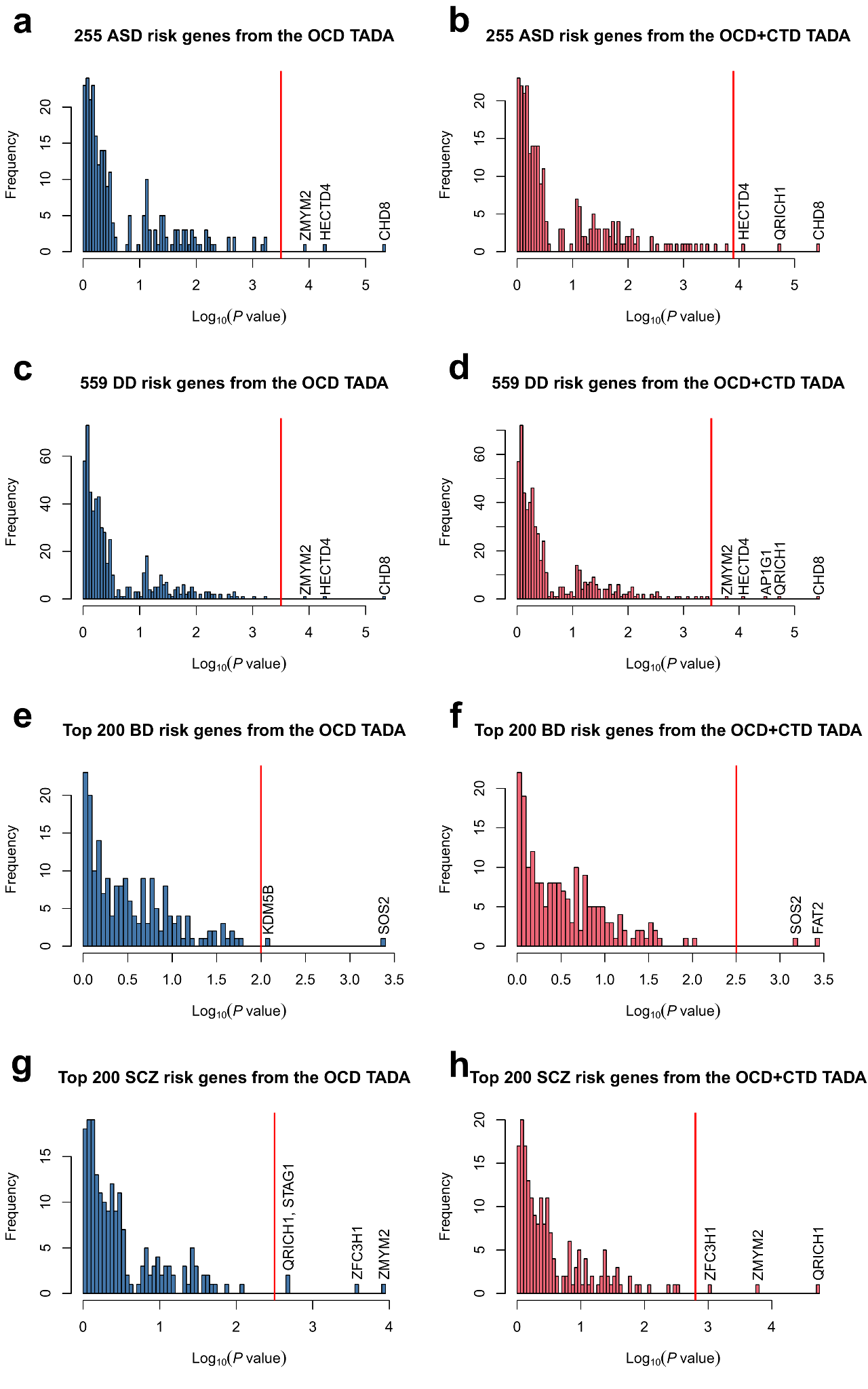

**eFigure 1**. **Distribution of –log_10_(*P* value) of ASD, DD, BD and SCZ risk genes in the OCD and OCD+CTD TADA result.** **a,b)** 255 autism spectrum disorder (ASD) risk genes with q value<0.1 from Fu et al., 2022. **c,d)** 559 developmental delay (DD) risk genes with q value <0.1 from Fu et al., 2022. **e,f)** Top 200 bipolar disorder (BD) genes from Palmer et al. 2022 with Bipolar Exomes Browser (BipEx) study. **g,h)** Top 200 schizophrenia (SCZ) genes from Singh et al. 2022 with Schizophrenia Exome Sequencing Meta-analysis (SCHEMA) study. In each panel, relevant genes above the given threshold (vertical red line) are labeled.

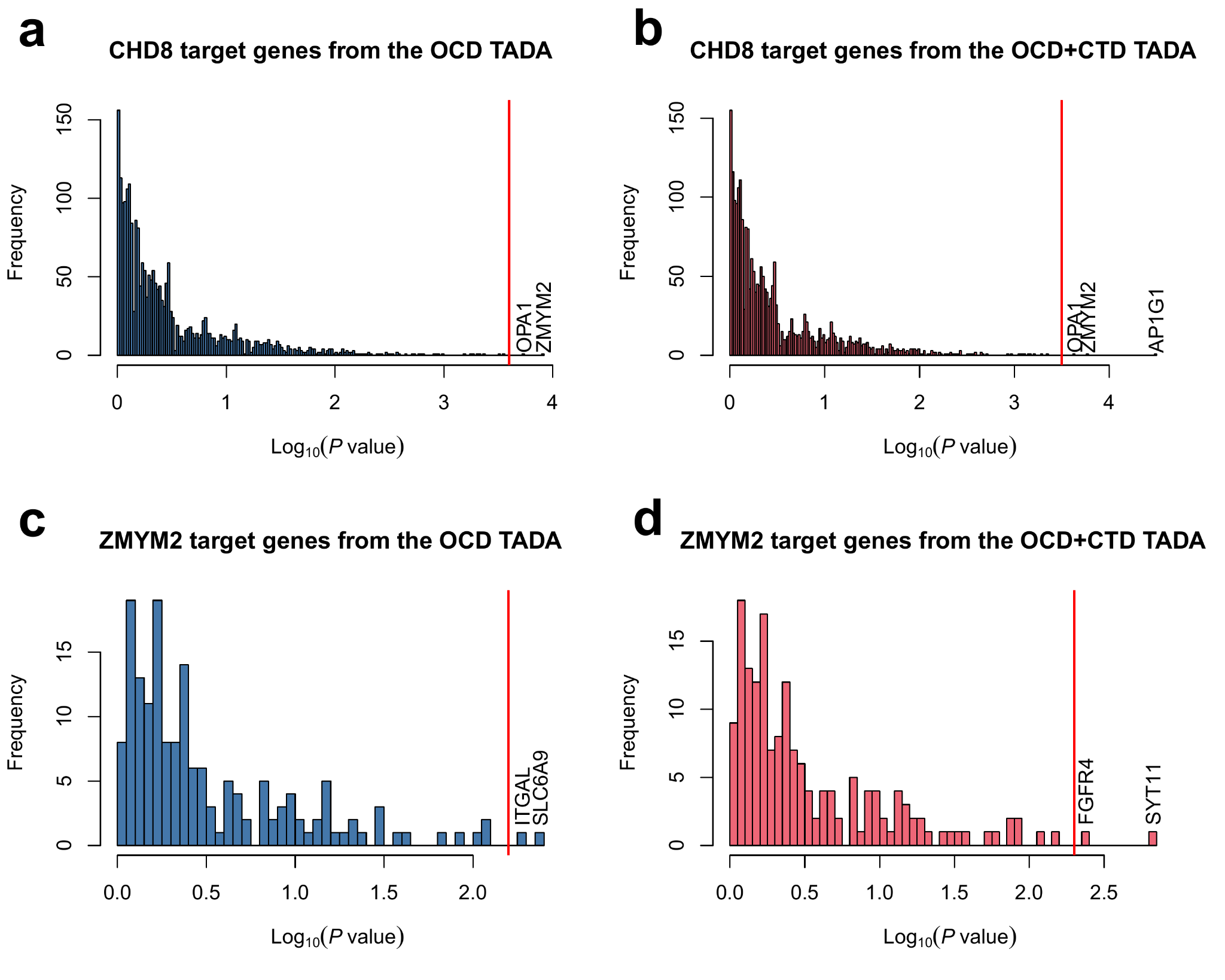

**eFigure 2**. **Distribution of –log_10_(*P* value) of CHD8 and ZMYM2 target genes in the OCD and OCD+CTD TADA result.** **a,b)** 2,327 CHD8 target genes (ChIP-seq analysis in hNSC&Brain from Cotney et al., 2015). **c,d)** 168 ZMYM2 target genes (overlapping genes from DEG analysis from Owen et al., 2023, DEG analysis from Graham-paquin et al., 2023, DEG analysis from Lezmi et al., 2020, and DEG analysis from Tsusaka et al., 2020). In each panel, relevant genes above the given threshold (vertical red line) are labeled.

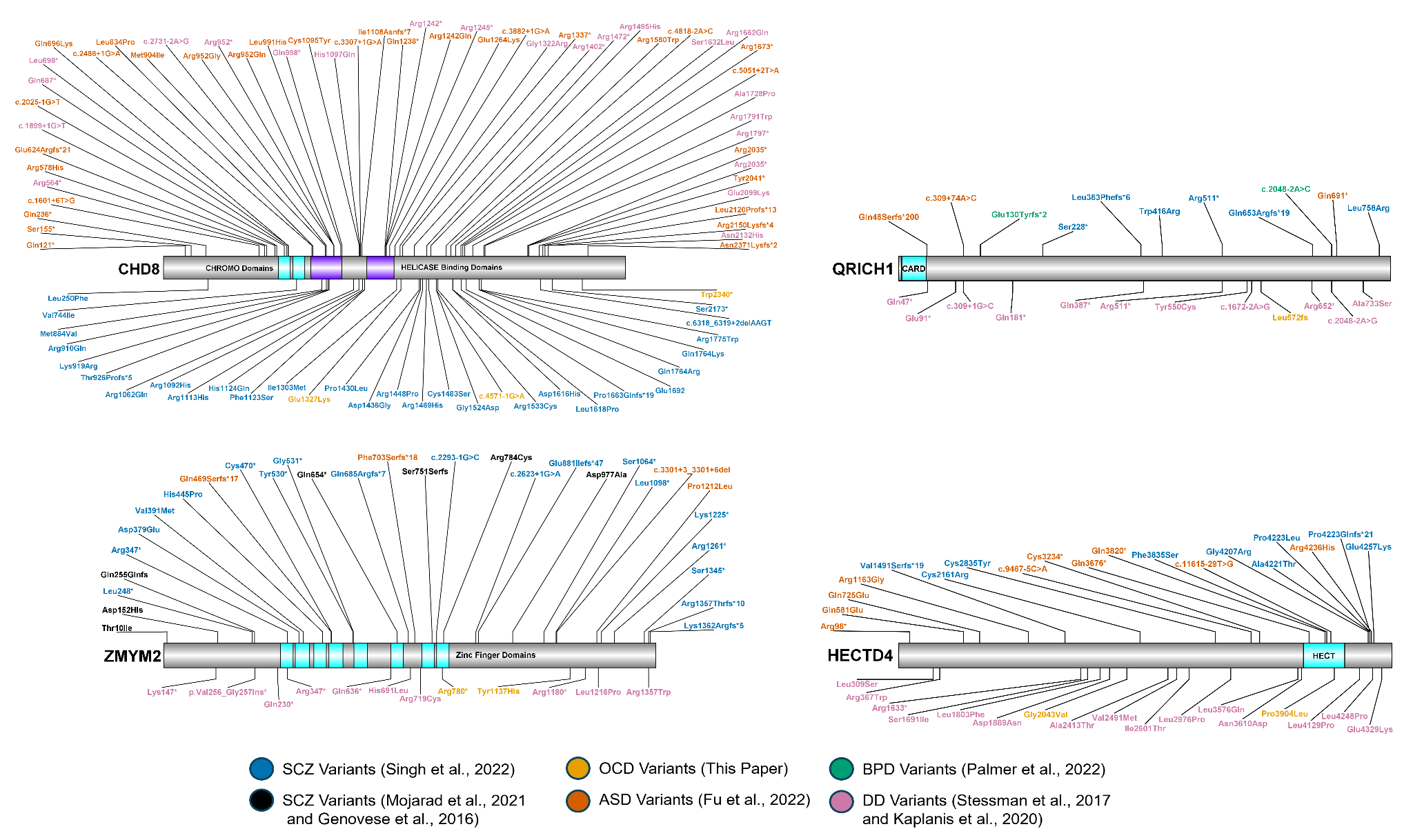

**eFigure 3. Schemes showing variants in CHD8, HECT4, QRICH1, and ZMYM2, identified in five psychiatric phenotypes.** Variants reported in the indicated papers are shown.

ASD, autism spectrum disorder; BD, bipolar disorder; DD, developmental delay; OCD, obsessive-compulsive disorder; SCZ, schizophrenia.
